# Music Medicine and Music Therapy in Pediatric Care: A systematic review of passive music listening research applications and findings on infant development and medical practice

**DOI:** 10.1101/2024.04.02.24305202

**Authors:** Efthymios Papatzikis, Maria Agapaki, Rosari Naveena Selvan, Deanna Hanson-Abromeit, Christian Gold, Shulammit Epstein, U Wun Vivian Lok, Evrykleia Barda, Varun Pandey

**Author notes:** Corresponding author (EP).

## Abstract

In recent years, the use of music as a therapeutic and developmental tool for infants, especially within neonatal intensive care units (NICUs), has seen a surge in interest. Despite a growing body of research underscoring the potential benefits of music therapy and music medicine in enhancing infant development and aiding medical practices, the specific characteristics of music that maximize these benefits remain poorly understood. This systematic review aims to fill this gap by investigating the effects of passive music listening on the development and medical outcomes of infants, both full-term and premature. Following the PRISMA guidelines, a comprehensive literature search was conducted, covering studies published up until December 2022. The focus was on passive music listening, with a deliberate exclusion of active music interventions. Out of the initial pool of studies, 56 met the inclusion criteria, determined by the PICO framework, focusing on passive music exposure among full-term and preterm infants. Starting with a descriptive analysis approach, the study employed Non-negative Matrix Factorization (NMF) and Latent Dirichlet Allocation (LDA) to identify key themes, including the physiological impacts of music, its role in pain management, effects on sleep and stress, and influences on feeding and weight gain. The review revealed a predominance of quantitative research methods, a significant concentration of studies from the United States, suggesting potential geographical bias, and a notable clinical setting bias. These findings indicate a critical need for methodological diversity and a more culturally inclusive and interdisciplinary approach to research. Although this systematic review highlights the beneficial role of passive music listening in pediatric care, it also points to the necessity for standardized music intervention protocols to optimize therapeutic and developmental outcomes for this vulnerable population. Future research should aim to bridge the methodological gaps identified, integrating both qualitative and quantitative approaches to provide a comprehensive understanding of music’s impact on infant development and medical practices in a global, culturally nuanced context.

## Introduction

Recent years have witnessed a surge in interest concerning the employment of music in infant cribs, and even incubators within neonatal intensive care units (NICUs). This increasing attention stems from a multitude of studies establishing the potential advantages of music therapy and music medicine for infant development, as well as its contributions to medical practices. Remarkably, infants exhibit a heightened responsiveness to auditory stimuli, which is attributed to their auditory system’s early maturation. Even before birth, they can perceive and process complex auditory inputs [8, 29], while this precocious auditory sensitivity is thought to play a pivotal role in the subsequent development of an array of cognitive and socio-emotional skills. Infants exposed to music early on have been observed to display improvements in cognitive, motor, and social-emotional development [3, 13, 44, 5], crediting for example, music interventions with augmenting infants’ attention, memory, and problem-solving capacities [45]. Additionally, music has been shown to bolster motor skills, like coordination and rhythm synchronization, by furnishing a structured, temporally organized context for movement [17, 18, 30, 36].

Music, and lullabies more specifically, have been shown to foster social-emotional development in infants. Activities involving singing and musical interaction with caregivers can enhance attachment and bonding while also promoting empathy, emotional regulation, and social communication skills [41, 22, 43]. Furthermore, the calming and soothing effects of lullabies are well-established in both premature and healthy infants, showing to impact medical practices, including pain management, stress reduction, and sleep promotion [20, 4, 38, 26, 6, 11, 35].

Yet, the complexity of music’s impact on infant development, especially in preterm infants, remains an area fraught with ambiguities and methodological discrepancies. This is starkly highlighted in the recent Cochrane review by Haslbeck et al. [16], which analyzed 25 trials with over 1500 infants and parents in NICUs. Their findings paint a picture of inconsistent outcomes and methodological diversity, failing to establish a clear consensus on the efficacy of music therapy and music medicine in this context. Specifically, they found no significant improvements in oxygen saturation or developmental scores and only minimal impact on parental anxiety, contrasting with the expected benefits often highlighted in literature. Although a reduction in heart rates of infants was indeed observed, this singular positive outcome amidst a sea of ambiguous or non-significant results calls into question the overall effectiveness and application of such interventions in NICUs.

As a result, one overarching observation in this realm of research is the existence of a significant knowledge gap regarding the specific characteristics of music - or more precisely, the design of music delivery protocols based on specific characteristics of music - that optimize its developmental and medical benefits and may be particularly - but not exclusively - pronounced in the context of neonatal intensive care units (NICUs) where passive listening [as opposed to active listening; for examples see 13, 32, 19, 12] could be a key intervention for vulnerable infants (please see below for more information on this critical yet existing dichotomy in this context). This gap, as evidenced by the findings of Haslbeck et al. [15], leads to the current study’s endeavor to systematically review the literature on the framework and impact of passive music listening on infant development and medical practice.

Nevertheless, while previous reviews [for example, please see 4, 10] have predominantly focused on ‘how’ music influences these areas, the present study aims to extend this understanding by addressing the ‘whats’ of the existing literature. This involves a comprehensive examination of what is currently known and mostly practiced at a technical and research design level for example, what are the groups of infants that are mostly studied, what are the dominant methodological trends and analytical techniques used in these studies, what are the major settings and lengths of these studies, what interdisciplinary approaches are involved, what key themes and patterns exist across these studies, and what areas are perhaps under-researched or lack sufficient data, to name just a few. Such an approach is critical for building a complete picture of the field, identifying the strengths and limitations of current knowledge, helping us to finally explore in a more comprehensive and practical way the generic research question: ‘How does passive listening of music influence medical practices and development in infants, both premature and full-term’?

By focusing on answering this question through the aforementioned angle, this study aspires to contribute to a more nuanced understanding of the role of passive music listening in infant care, aims to bridge the gap between theoretical knowledge and practical application through a different point of access to evidence, while guiding future research and clinical practices to optimize the use of music therapy and music medicine as developmental and therapeutic tools for infants. The objective is to provide actionable insights that can inform policies and protocols in this very sensitive yet extremely interdisciplinary context, ultimately benefiting this vulnerable population in a more informed and effective manner.

### Why Passive Music Listening?

This review specifically concentrates on the effects of passive music listening on infant development and medical practice. For that matter, we approach passive listening as entailing infants being exposed to calming, soothing music or lullabies without active engagement or interaction with musical stimuli or caregivers. This form of listening primarily engages the auditory system, in contrast to active listening, which involves direct engagement with musical stimuli, often through interactive activities with caregivers or participation in musical games, engaging multiple sensory pathways including tactile and visual senses. In the context of neurodevelopmental impact, the study by Remijn and Kojima [32] sheds light on the distinct neural processing involved in passive versus active listening. This contrast between passive and active music listening modes is essential to understand, as they may influence infant development via separate neural pathways and cognitive processes, while especially for preterm infants in Neonatal Intensive Care Units (NICUs), where active engagement is not always possible, passive music listening may serve as the only practical approach for providing auditory enrichment, while offering a soothing and stabilizing effect for infants in resting states or during periods of restricted caregiver interaction.

### Methodology

#### Literature review design

In compliance with the PRISMA (Preferred Reporting Items for Systematic Reviews and Meta-Analyses) guidelines [25], we conducted a comprehensive and systematic literature search. Our search covered the Google Scholar database for pertinent articles published up until December 2022. We opted to use only Google Scholar for our systematic review due to its strengths in providing comprehensive cross-disciplinary results, expansive coverage of both scientific journals and gray literature. This approach proved to be particularly suitable for our topic, which intersects with various fields including music therapy, music medicine, clinical care, pediatrics, neurology and of course music, ensuring we did not overlook any relevant studies that might not be indexed in more specialized databases like PubMed. Furthermore, we also checked manually 3 published systematic reviews [24, 31, 42] which were close or related to the domain pertaining to our study.

In our search approach, we employed search terms (in variable sequence) such as ‘lullaby’, ‘music’, ‘crib’, ‘brain’, ‘development’, ‘infant’, and ‘cradle’, intentionally excluding context or framework specific terminology such as ‘music therapy’, ‘music medicine’, ‘pediatric’ or ‘clinical’. This choice was predicated on the understanding that the former broader search terms would inherently capture relevant information or nuances from the latter more focused on context or framework areas without restricting our review scope. In order to qualify for inclusion in this systematic review, studies had to meet the following PICO framework criteria:

- Populations: full-term and/or preterm infants
- Interventions: passive listening of music or lullabies
- Comparisons: not applicable
- Outcomes: medical treatment, biological development and/or clinical contexts

Furthermore, studies had to be in English and published following a peer-reviewed procedure, while all of them had to be original studies (Randomized/Non-randomized). We excluded studies based on the following exclusion criteria:

- *Mutli-modal Interventions*: Studies employing multi-modal or multisensory protocols offer information on interventions in mixed ways with music, making it difficult to isolate the effects of music from other sensory applications.
- *Outcomes*: Studies primarily centered on cultural, cognitive, or musicological aspects of infant music exposure.
- *Study types*: Review articles or meta-analyses were not included in the analysis, but reference lists of such reviews were examined for potentially relevant original studies

#### Study selection

Two independent reviewers were tasked with screening the titles and abstracts of the retrieved articles using the aforementioned eligibility criteria. The same reviewers then assessed the full-text articles that satisfied the initial screening criteria for eligibility. In cases where disagreements arose between the reviewers, a consensus was reached through discussion, or by consulting a third reviewer, if necessary.

#### Study Sample

The initial search identified a total of 1,042 articles. After removing duplicates and checking for relevance, 146 articles remained for an initial abstract screening. Following this initial screening, 104 articles were considered for full-text review. Of these, 56 articles met the eligibility criteria and were finally included in the systematic review. A flow diagram of the study selection process is presented in Figure 1.

**Figure 1.**
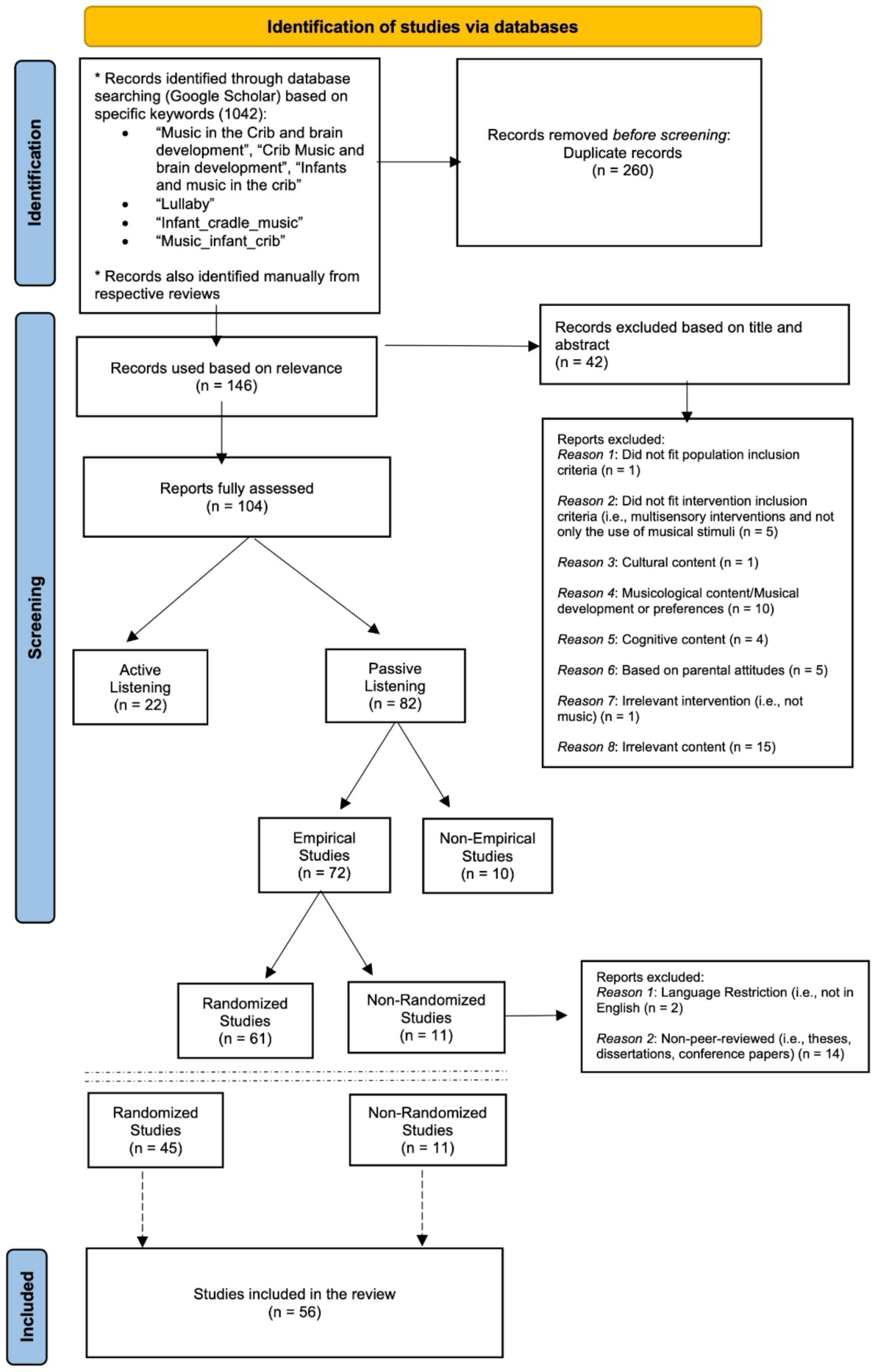
Flow Diagram of the study selection process.

#### Data Extraction

Before beginning the data extraction process, two custom Excel spreadsheets were designed to serve as data extraction tools. The first spreadsheet featured 23 primary headings, each divided into multiple subcategories [see supplementary file SF1 for specifics], while the second spreadsheet was designed to catalog the key findings, strengths, and limitations of each one of the 56 studies [see supplementary file SF2 for specifics]. In the first spreadsheet, the headings were carefully selected to capture a broad range of variables including bibliographic details, research methodologies, analytical approaches, and demographic information. With this tool, the following details were extracted and further used for a descriptive statistics analysis:

- Bibliographic Information: This included the title, number of authors, country of origin, and publication year.
- Research Focus and Context: The primary knowledge context (Development, Medical/Treatment/Therapy, Prevention) was identified.
- Methodology and Quality: The research methods employed were categorized as qualitative, quantitative, or mixed. Information on the validity and reliability of these methods was also noted.
- Analytical Techniques: The types of qualitative and quantitative analyses used were extracted.
- Data Collection: Information about how the data were collected and which tools were used was documented.
- Sample Characteristics: This included the population focus (term newborns, preterm newborns, infants) and any pathological conditions mentioned.
- Research Context: The setting in which the research was conducted was noted, such as a hospital, nursery, or domestic environment.
- Length of Study: This was captured to understand the time frame over which observations were made.
- Interdisciplinary Involvement: Any involvement from other disciplines or professions was extracted.
- Gender and Sex Orientation: Information was extracted on whether the study separated data by gender or specified the sexual orientation of the sample.

To ensure the reliability of the data extraction process, a ‘double-extraction’ method was employed, where two independent researchers extracted data from the same article. Any discrepancies were resolved through discussion or consultation with a third researcher.

Furthermore, to deeper understand the content of our study sample, a systematic analysis involving just the findings of all 56 studies was also undertaken. Using in this case the second spreadsheet as our data extraction tool, we performed a meticulous review of each article to export and organize their findings individually. This spreadsheet served as a foundational dataset, on which a subsequent Non-negative Matrix Factorization (NMF) was performed (using a custom made Python code), as well as a Latent Dirichlet Allocation (LDA) for topic modeling and in-depth analysis of emerging themes and patterns within the specific research landscape.

The choice of NMF was motivated by the several advantages it offers, including its capacity to generate interpretable topics, computational simplicity, and sparse representation. NMF allows for a minimal number of topics to describe each available dataset, facilitating easier interpretation and meaningful topic discovery. On the other hand, LDA, while simpler and less powerful than NMF, played a complementary role in our text analysis. It served to cross-reference the results obtained from NMF, ultimately providing an averaged computation of the most prominently emerging topics within the findings of the 56 reviewed studies.

The NMF technique involved the conversion of the textual data into a numerical representation using Term Frequency-Inverse Document Frequency (TF-IDF) vectorization. The TF-IDF is a numerical statistic used in text mining and information retrieval to reflect how important a word is to a document in a collection or corpus of documents (dataset). It is a way to weigh the importance of terms (words) in the document based on how frequently they appear across all documents, while it is calculated with the following equation:

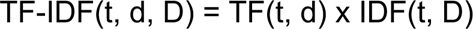

Where t = Number of times term t appears in document/text d

Where d = Total number of terms in document/text d

Where D = Total number of documents/text in corpus D

After the TF-IDF matrix generated (see SF3), it was then decomposed into two lower-dimensional matrices, *W* (see SF4) and *H* (see SF5); where *W* represents the “document-topic” relationship and *H* encapsulates the “topic-word” relationship. The *H* matrix was examined to identify the top words that are representative of each topic, and then each “document” (in our case, each study’s key findings) was assigned to the topic for which it had the highest representation in the *W* matrix.

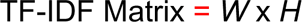

In contrast to the TF-IDF-based approach used for NMF, LDA operates by modeling documents as mixtures of topics, where each topic is characterized by a distribution of words. In our LDA analysis, the textual data underwent preprocessing, including tokenization and stop word removal. LDA then inferred the underlying topics within the corpus of documents (dataset) and the distribution of words associated with each topic. This process involves a probabilistic model that assigns words in each document to specific topics, and topics to documents, aiming to find a coherent and meaningful representation of the content. After running LDA, we obtained a set of topics and their associated word distributions. These topics served as a way to categorize and summarize the content of the reviewed studies. For each topic, we identified the most representative words, offering insights into the key themes present in the dataset.

Following the completion of both NMF and LDA analyses and the identification of primary topics within the key findings of the 56 studies, Principal Component Analysis (PCA) emerged as a final yet critical step in our research methodology. The utilization of PCA served several pivotal functions in enhancing our understanding of the findings dataset and further refining our analytical approach. Initially, NMF and LDA provided valuable insights into the specific dataset by identifying key topics and recurring themes. However, the inherent complexity of the dataset, consisting of numerous variables and topics, required a means of dimensionality reduction to simplify interpretation. This necessity led us to incorporate PCA into our analysis. In that sense, PCA effectively reduced the dimensionality of the high-dimensional dataset, while this transformation facilitated a more concise and comprehensible representation of our dataset, enabling us to delve deeper into the underlying patterns of the findings. As PCA allowed us to visualize the intricate relationships and dependencies between topics and variables, it provided a visual context that was complementary to the results obtained from NMF and LDA.

## Results

### Descriptive Statistics

Our analysis rigorously assessed the extensive collection of studies carried out between 1990 and 2022 (Figure 2). Though the studies were geographically diverse, a significant portion originated from the United States (Figure 3). In terms of authorship, the studies exhibit considerable variation, with the number of contributing authors ranging from a minimum of one to a maximum of 13 (Figure 4).

**Figure 2.**
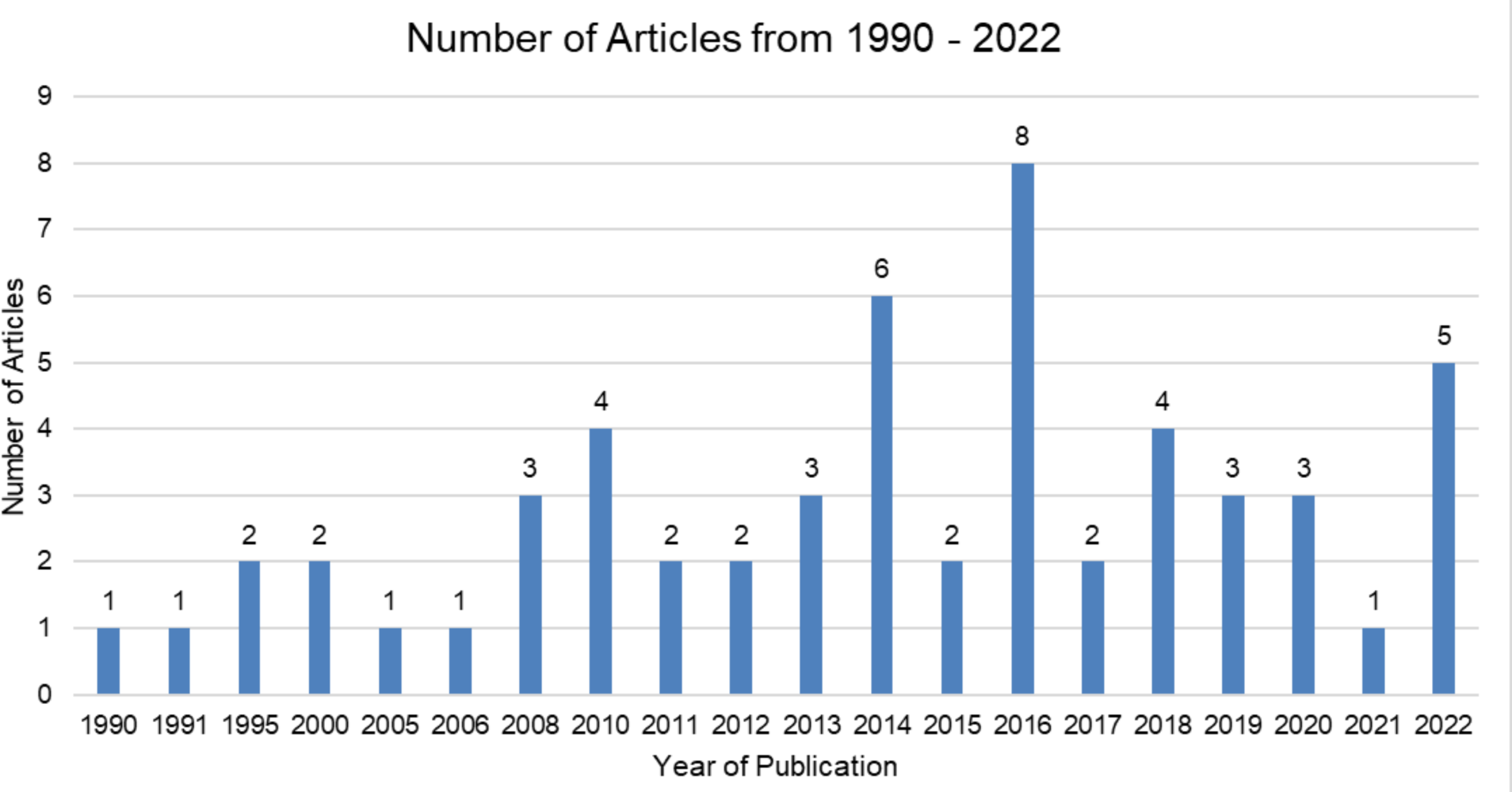
Studies conducted between 1990 and 2022 per year.

**Figure 3.**
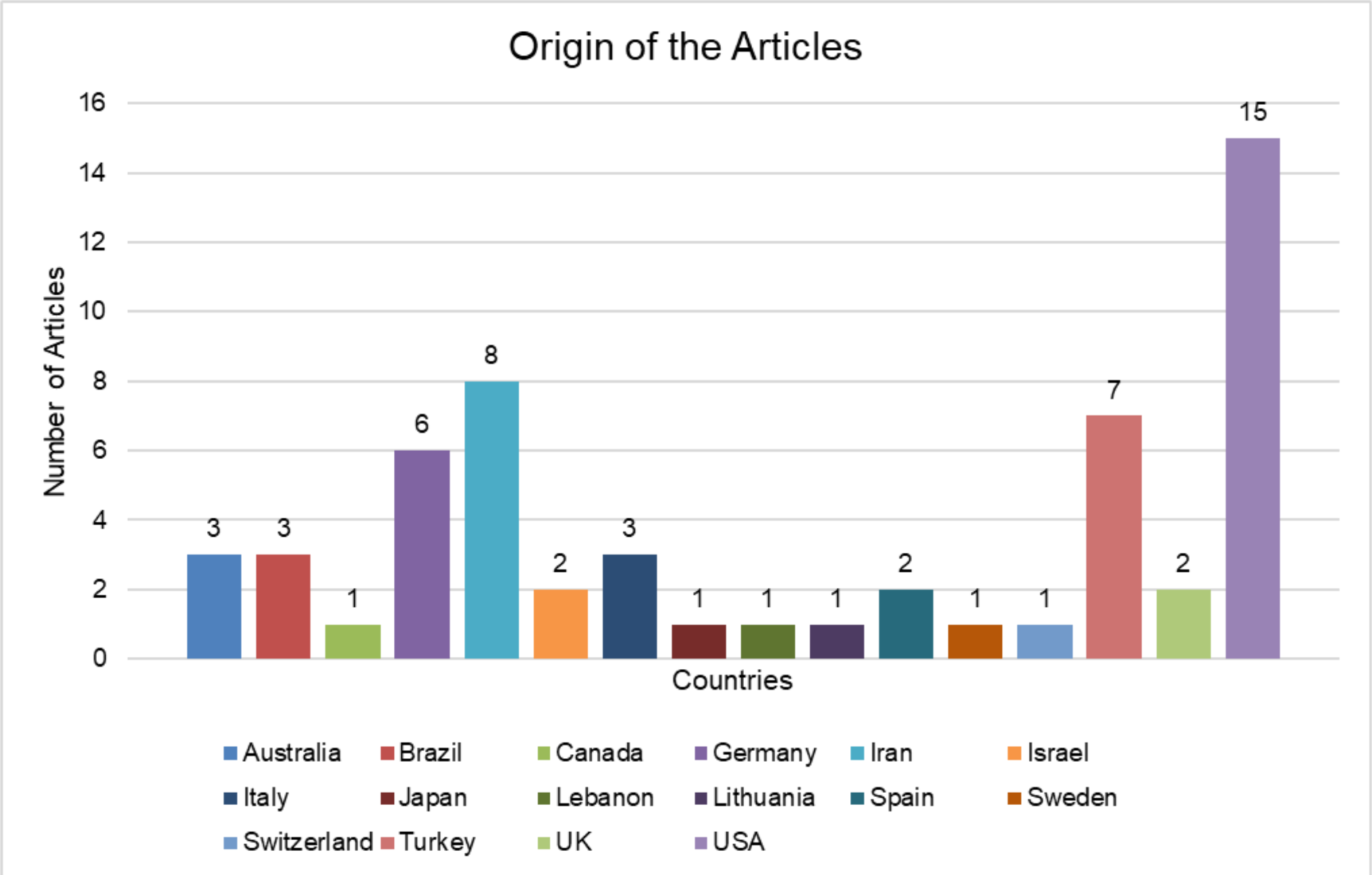
Geographical origin of the studies carried out between 1990 and 2022.

**Figure 4.**
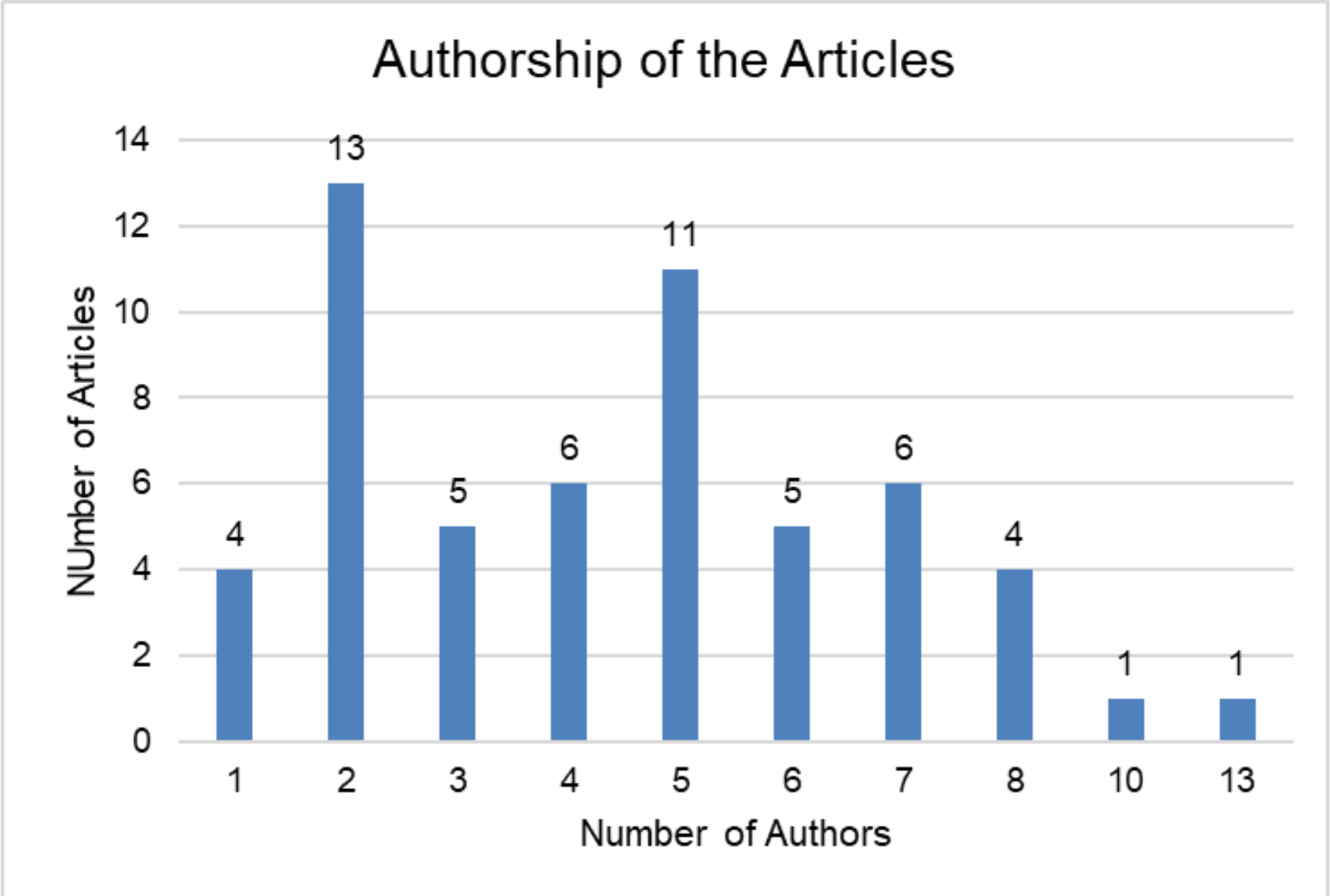
Contributing authors per article.

#### Thematic analysis of titles

An analysis of the study titles - using a custom made python script - revealed the most researched broader themes (Table 1), which included the influence of mothers, indicated by words such as “mothers” and “maternal”; the timing of interventions or observations, represented by the word “during”; and the use of recorded music in studies, implied by the term “recorded”. Other focal points were pain management, physiological focus, therapeutic methods, and a comparison between live and recorded interventions or stimuli, highlighted by words like “pain”, “physiological”, “responses”, “therapy”, “live”, and “singing”. This thematic analysis was derived from a word frequency count on the study titles, excluding common English words like “the”, “and”, “of”, “in”, and “to”, which do not contribute to the thematic content.

**Table 1.**
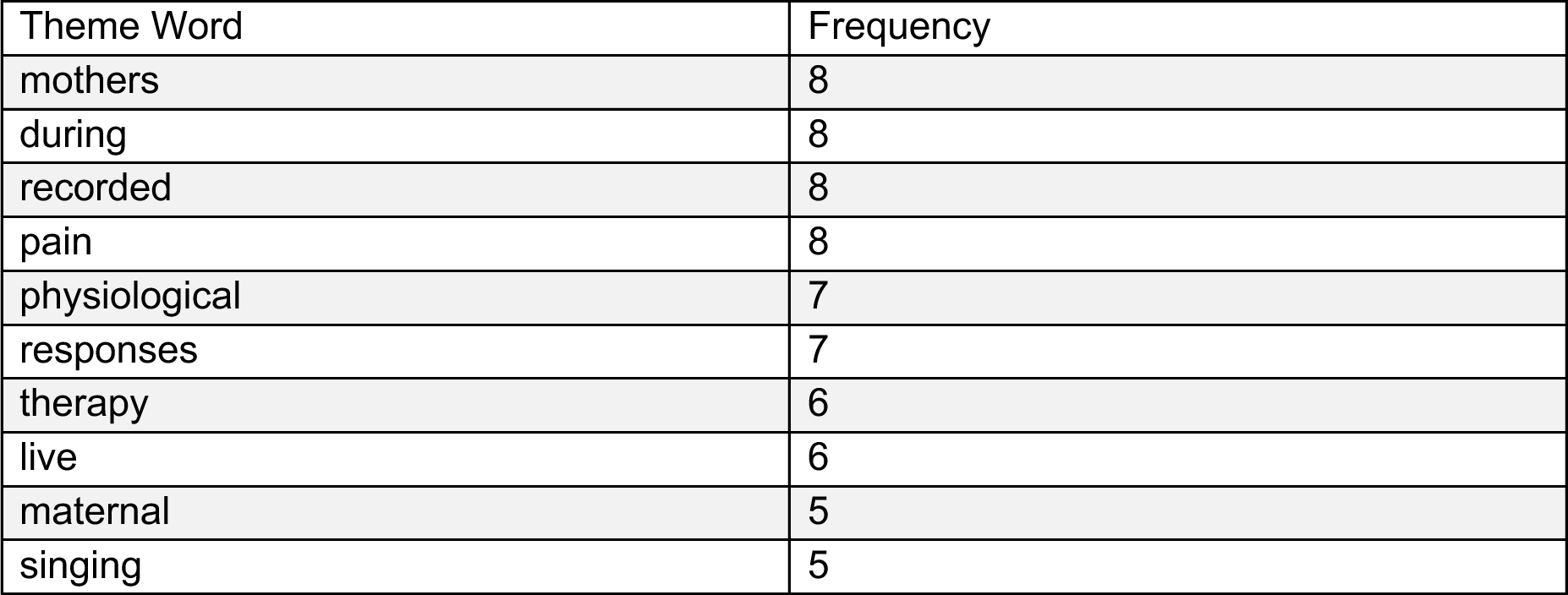
Frequency of the most researched broader themes in study titles.

| Theme Word | Frequency |
| --- | --- |
| mothers | 8 |
| during | 8 |
| recorded | 8 |
| pain | 8 |
| physiological | 7 |
| responses | 7 |
| therapy | 6 |
| live | 6 |
| maternal | 5 |
| singing | 5 |

Further, a co-occurrence analysis of theme-related words in the study titles brought to light interesting relationships and potential focal points in the research landscape. For instance, the terms “maternal” and “singing” co-occurred four times, suggesting a significant focus on the influence of maternal singing on various outcomes, possibly including infant development or psychological well-being (see Table 2). Similarly, the pairing of “physiological” and “responses,” appearing together four times, indicated a frequent exploration of physiological responses to different stimuli or conditions. The analysis unveiled several other notable co-occurrences, such as “during” and “singing” and “during” and “pain”, each appearing three times, hinting at studies focusing on real-time effects of singing and pain management techniques during specific procedures or behavior states, respectively. This detailed co-occurrence analysis not only helped us underscore the interrelated themes in the dataset but also pointed out a quite good profile of diversity of research avenues explored in the studies, ranging from the therapeutic effects of singing to the comparative analysis of live versus recorded stimuli.

**Table 2.** A co-occurrence analysis of theme-related words in the study titles.

| Co-occurring Themes | Frequency |
| --- | --- |
| ('maternal', 'singing') | 4 |
| ('physiological', 'responses') | 4 |
| ('during', 'singing') | 3 |
| ('during', 'pain') | 3 |
| ('during', 'maternal') | 2 |
| ('maternal', 'recorded') | 2 |
| ('live', 'recorded') | 2 |
| ('live', 'physiological') | 2 |
| ('singing', 'therapy') | 2 |
| ('physiological', 'recorded') | 1 |
| ('recorded', 'responses') | 1 |
| ('live', 'responses') | 1 |
| ('recorded', 'therapy') | 1 |
| ('recorded', 'singing') | 1 |
| ('maternal', 'therapy') | 1 |
| ('during', 'physiological') | 1 |
| ('pain', 'physiological') | 1 |
| ('during', 'therapy') | 1 |
| ('during', 'live') | 1 |
| ('live', 'therapy') | 1 |
| ('live', 'singing') | 1 |
| ('during', 'mothers') | 1 |
| ('mothers', 'pain') | 1 |
| ('mothers', 'physiological') | 1 |
| ('live', 'mothers') | 1 |
| ('maternal', 'responses') | 1 |
| ('responses', 'singing') | 1 |

#### Quantitative vs Qualitative Approach

Out of the 56 studies analyzed, a significant portion adopted quantitative approaches (82%), while a smaller number were based on mixed (16%) and qualitative (2%) methodologies. Especially for the qualitative methods, we found a significant Chi-Square statistic (χ^2^ = 29.42) with a very low p-value (*p* = 0.0064) when examining the relationship between reported trustworthiness and existing credibility measures in qualitative methods (Figure 5). Furthermore, we found that the process of examining, interpreting and understanding non-numerical data collected during the studies were tested using five parameters namely constant comparison, thematic analysis, qualitative coding, analytic memos and discourse analysis (Figure 6).

**Figure 5.**
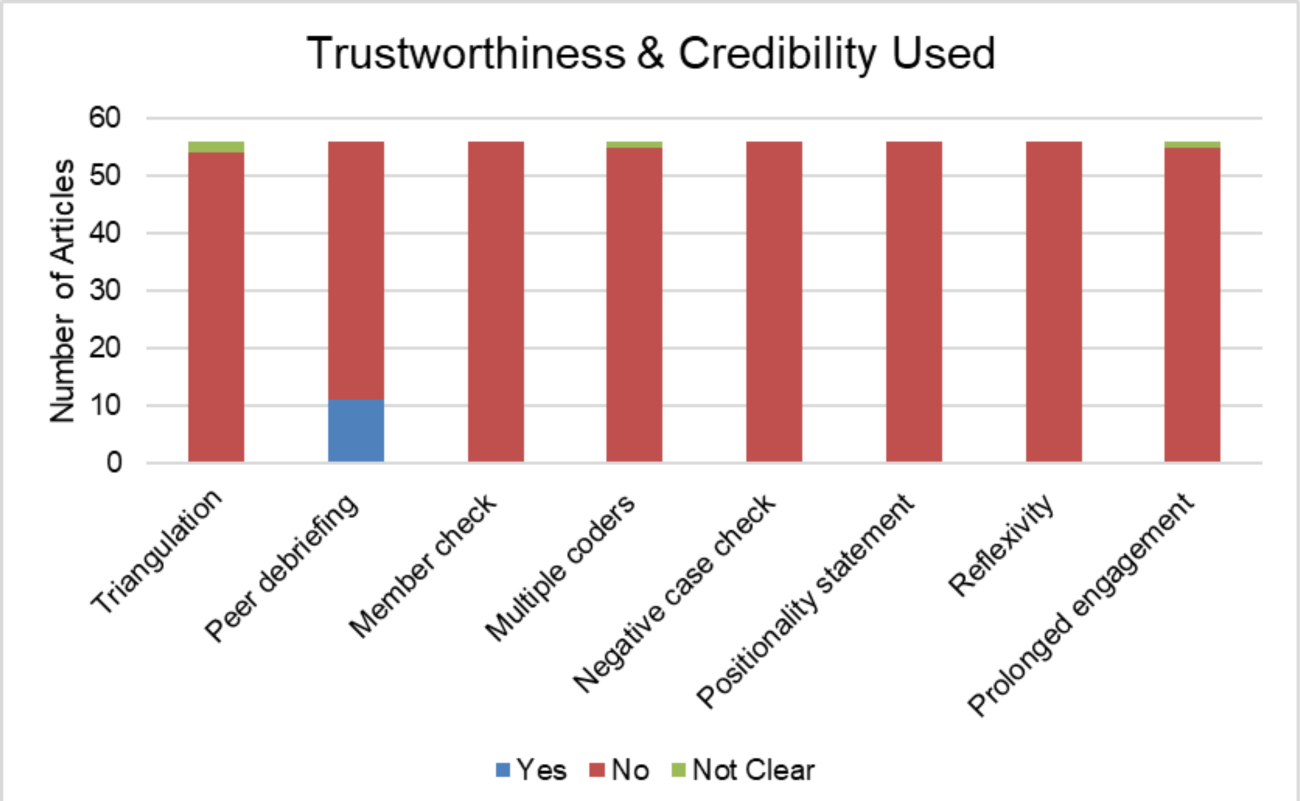
Relationship between reported trustworthiness and existing credibility measures in qualitative methods.

**Figure 6.**
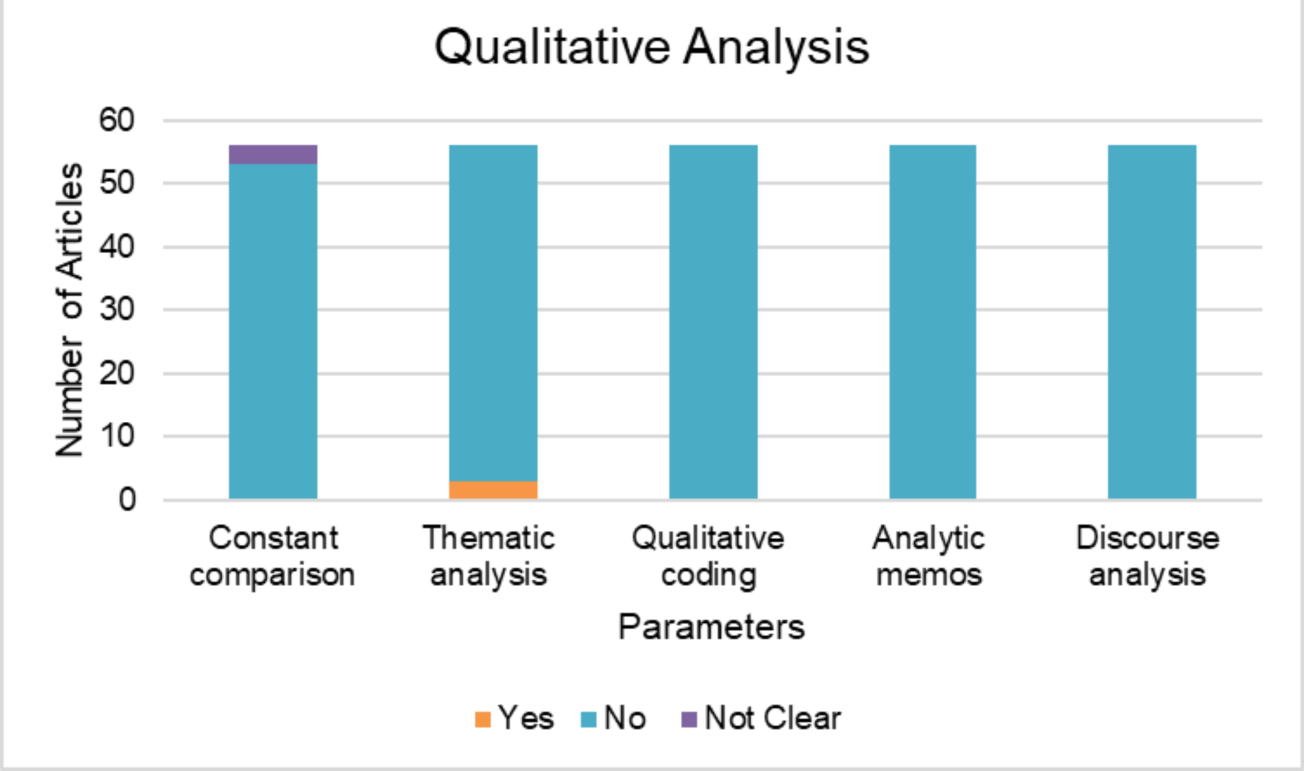
Parameters evaluated for the process of examining, interpreting, and understanding qualitative data.

On the other hand, for the quantitative methods, their statistical analyses predominantly adopted descriptive, testing of means, and correlational approaches (Figure 7) revealing a troubling lack of diversity. This was substantiated by a significant Chi-Square statistic (χ^2^ = 56.0) and an extremely low p-value (p = 0.007) shown after a correlational analysis of validity measures within these quantitative methods.

**Figure 7.**
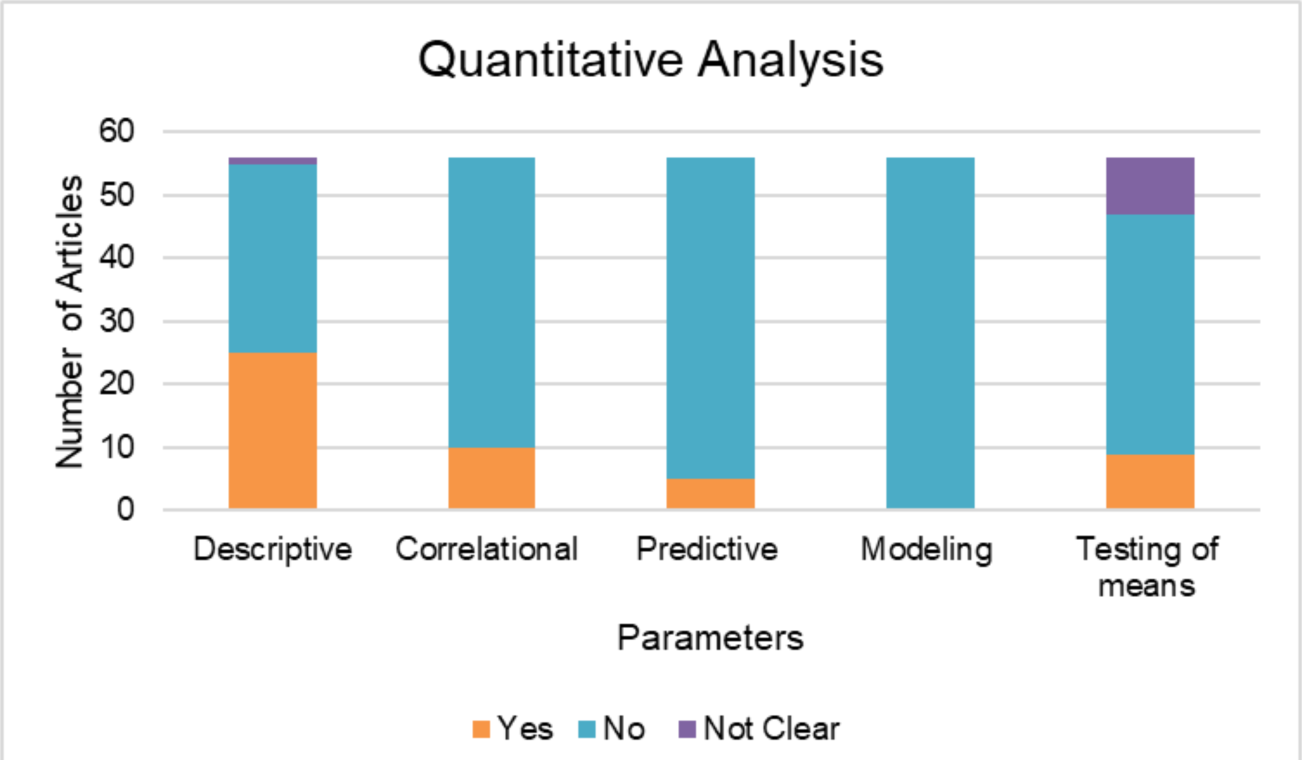
Statistical analyses used for the process of examining, interpreting, and understanding quantitative data.

#### Research Contexts

In the corpus of research studies analyzed, a predominant focus was observed on the application of music in medical treatment and therapeutic interventions, overshadowing its utilization in developmental and preventative contexts (Figure 8).

**Figure 8.**
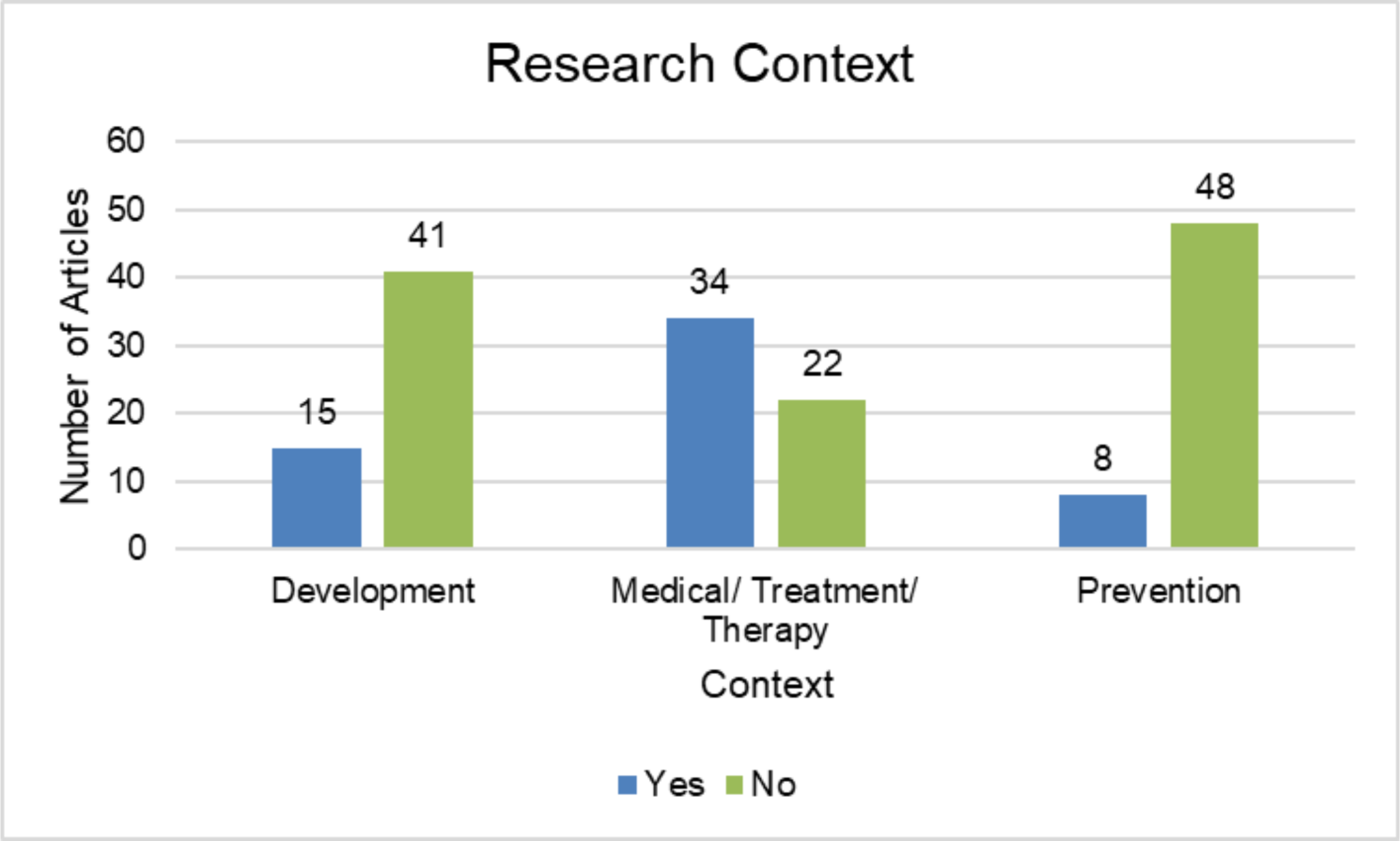
Research contexts used in selected studies.

#### Data collection methods

Our dataset revealed a diverse range of methods for data collection in these studies, encompassing physiological measures such as Heart Rate (HR), Oxygen Saturation, and Respiratory Rate (RR), psychological scales including the APGAR score and PIPP scale, and various questionnaires. Advanced neuroimaging techniques such as EEG, fMRI and LC-MS (Liquid Chromatography–Mass Spectrometry) were also utilized, alongside observation methods like interviews and video recordings. In total, there were 54 unique methods of data collection, each used in one study, except for fMRI and the PAL device, which were employed in two studies each.

#### Populations involved

In examining the populations studied, most studies focused on term newborns (11 studies) and preterm newborns (47 studies) either individually or in combination. Studies mentioning infants only - as a generic code - as their respective studied population were very rare, appearing only in one of the studies.

#### Setting of research

A striking 96% of the studies within this research domain were conducted in hospital settings, highlighting the field’s pronounced emphasis on clinical contexts. Conversely, non-clinical settings such as domestic environments and rehabilitation centers were significantly under-represented, constituting only 4% and 2% of the 56 studies, respectively. It should be noted that the cumulative percentages may exceed 100% due to the inclusion of multiple settings within individual studies.

#### Gender

The field of gender studies has adopted a varied approach; some studies have refrained from differentiating based on gender, while others have concentrated exclusively on one gender or have established distinct groups for male and female participants. The following figure (Figure 9) encapsulates the outcomes of our analysis with respect to the presence or absence of methodological differentiation based on gender. A total of 18 studies explicitly mentioned gender separation in their methodology, while 26 studies did not mention gender differentiation in the methodological section - yet they revealed such differentiation during the data analysis phase. Notably, one study utilized parents’ reflections as a method to ascertain the gender of the subjects. In the remaining 11 studies, gender separation was neither mentioned nor implemented.

**Figure 9.**
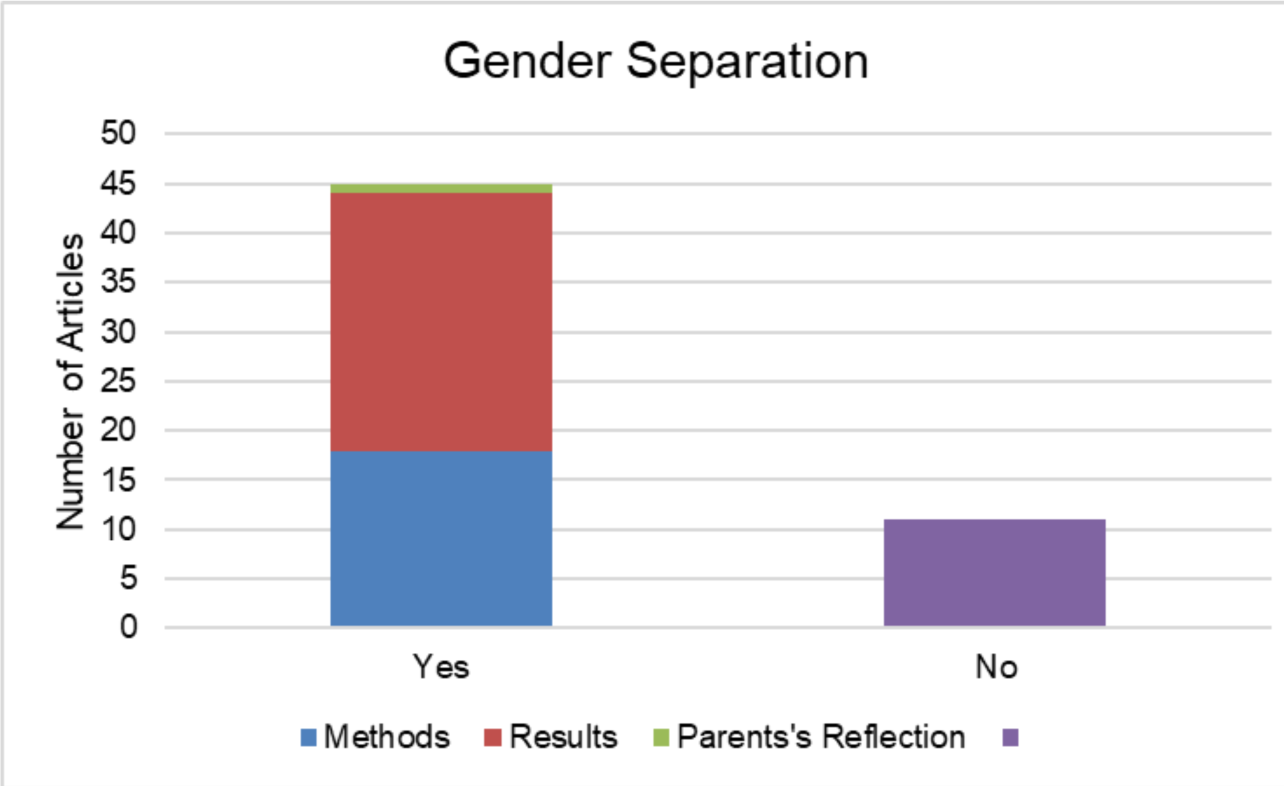
The presence or absence of methodological differentiation based on gender.

#### Research tools

In our scrutiny of the 56 studies, we noted the utilization of a broad spectrum of research tools employed across different methodologies and data collection strategies. These tools can be categorized into the types shown in the following table (Table 3):

**Table 3.** Research tools and their descriptions used in various techniques and data collection strategies.

| Category | Description |
| --- | --- |
| Audio Equipment | Includes micro-audio systems, speakers, headphones, and MP3 players. |
| Video Equipment | Various models of digital video cameras were used. |
| Medical Research Devices | Devices such as pulse oximeters, decibel meters, and thermal analyzers were common. |
| Tools for Clinical Analysis | These encompassed specialized software and devices for data analysis like ELISA, ActiGraph, and HRV Scanner. |
| Miscellaneous | This category captured custom-made tools, musical instruments, and other specialized equipment employed for unique study requirements. |

#### Representation of Professionals

Our analysis revealed a conspicuous lack of detail in depicting the range of professionals engaged in the 56 studies. These ranged from nurses and music therapists to audiologists and academic researchers, including both codes of “chief investigator” and “nurse researchers”. Importantly, when we examined the correlation between the disciplines or professions involved and the countries of origin, our statistical analysis yielded a strikingly high Chi-Square statistic (χ^2^ = 577.3) coupled with a remarkably low p-value (p = 0.003).

#### Journals-Pathology-Length of Intervention

In our comprehensive examination of the 56 studies, several journals emerged as dominant platforms for research in this specific domain. Notably, *Acta Paediatrica* and the Journal of Music Therapy each accounted for five publications, while the Journal of Neonatal-Perinatal Medicine, Pain Management Nursing, and Journal of Perinatology each contributed two publications. These figures highlight the significant role these journals play in shaping the academic discourse and advancing research in the field.

Turning to the issue of pathology, an analysis of the value distribution in our dataset points to an intriguing pattern. In 34 out of 56 studies, no pathology was explicitly mentioned, comprising approximately 61% of the research corpus. On the other hand, 22 studies did mention a pathology, making up around 39% of the studies.

Lastly, concerning the length of interventions, it is worth noting that this crucial aspect remains largely underreported. Out of the 56 studies, only two explicitly indicate the length of the interventions undertaken.

### Findings Analysis

Based on the results of both the NMF (see SF6) and LDA (see SF7) - i.e., their overlapping emerging themes - the main topics within the key findings of the 56 studies appeared to be the following (see also Figure 10 below for a numerical distribution):

- **Topic 0: Effects of Music on Infants’ Physiology**: Both NMF and LDA highlight the impact of different types of music (including lullabies and classical music) on infants’ heart rate, respiratory rate, and stress levels. This topic seems to be the most comprehensive, covering physiological responses to music interventions.
- **Topic 1: Pain Management**: Another common topic focuses on how music or sound interventions can alleviate pain, especially during procedures like glucose tests. Terms like ‘pain’ and ‘glucose’ frequently appear in the findings, often in the context of pain scores like PIPP (Premature Infant Pain Profile).
- **Topic 2: Sleep and Stress**: The role of music in affecting sleep quality and reducing cortisol levels in infants is another recurring theme. Both models identify terms related to sleep, stress, and cortisol levels.
- **Topic 3: Feeding and Weight Gain**: This topic centers on the effects of interventions on infants’ feeding habits and weight gain. Terms like ‘weight’, ‘feeding’, and ‘Non-Nutritive Sucking (NNS)’ appear in this context.
- **Topic 4: Methodological Aspects**: While not a ‘finding’ per se, the models also highlighted the frequent mention of study methodologies, with terms like ‘study’, ‘group’, and ‘therapy’ appearing often. This suggests that the dataset contains a variety of research designs and approaches.

**Figure 10.**
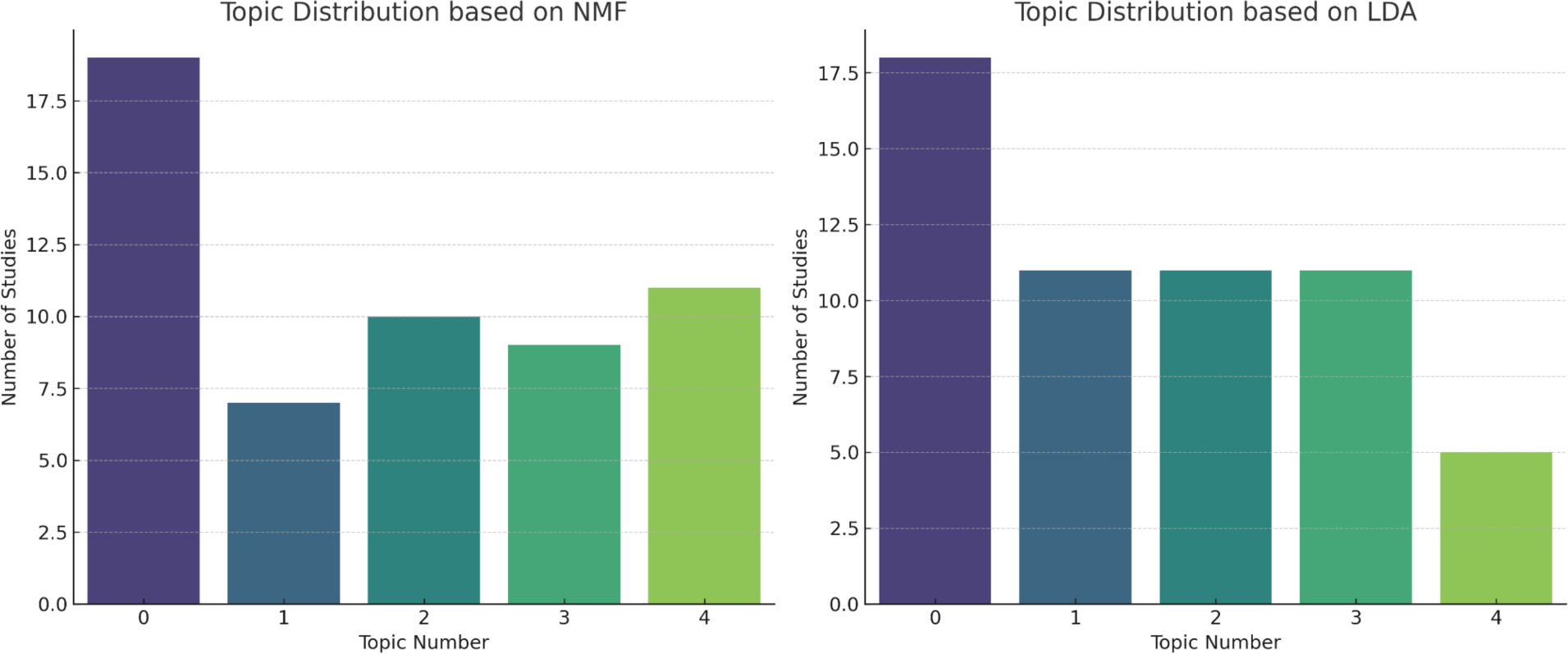
The numerical distribution of the main topics within the key findings of the 56 studies.

Finally, the application of the PCA significantly enhanced our understanding of the interrelationships among the key findings of this specific set of studies. By plotting the principal components against each other, we observed distinct clusters within the research landscape. As depicted in Figure 11, the studies are distributed across the PCA plot, indicating the multidimensionality of the research conducted in this field. Most notably, a particular cluster highlighted in red captured our attention due to its compactness. This cluster comprises studies with Study IDs: [8, 12, 23, 30, 44], which demonstrate a high degree of similarity in terms of their key findings. Such clustering within the PCA plot is indicative of a subset of research that, due to its consistency and potential cumulative evidence, may possess a higher degree of validity or reliability within the broader scope of our review’s thematic focus.

**Figure 11.**
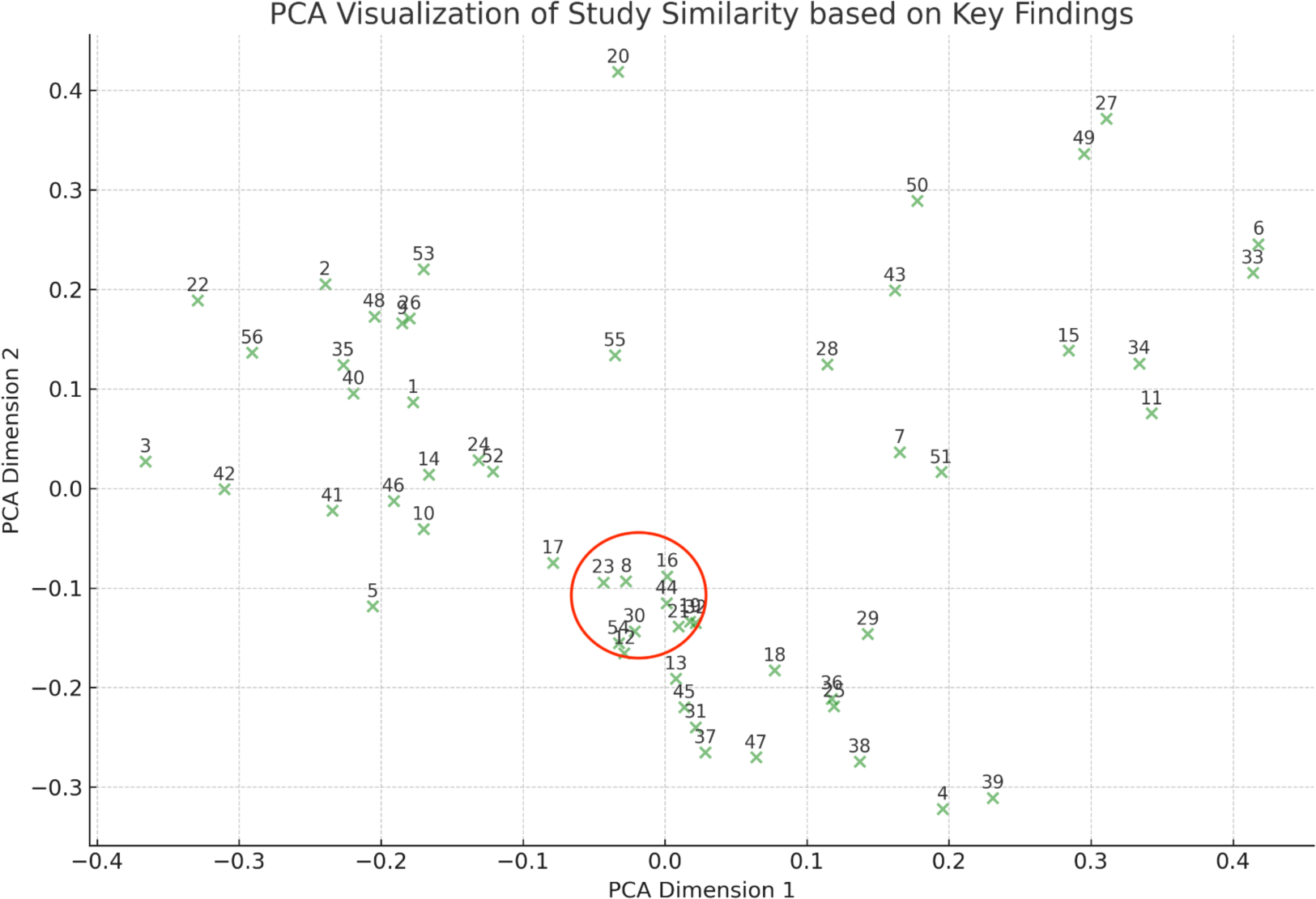
Interrelationships among the main findings of the 56 studies.

## Discussion

The increasing integration of music into the caregiving environment as a non-pharmacological tool reflects a burgeoning recognition of the former’s potential developmental and therapeutic benefits, while it also highlights its emerging importance in pediatric care. However, despite the growing body of research on this front, inconsistencies and a lack of consensus on the efficacy of music interventions in this context present the need for a more nuanced exploration of the former’s role in pediatric care. For this reason, adhering to PRISMA guidelines, we conducted a comprehensive and systematic literature search specifically on passive music listening, capturing a wide range of interdisciplinary studies until December 2022, ensuring an exhaustive inclusion of relevant literature spanning music therapy, music medicine, clinical care, pediatrics, and neurology. In total, 56 articles met our inclusion criteria, while provided a robust dataset to analyze, helping us to address the critical gap in the literature centered around the interplay between theoretical knowledge and practical application in this context. We achieved this by focusing more on the ‘whats’ of this link rather the ‘hows’, following a descriptive statistics comprehensive mapping of the studies’ profiles and methodological structuring, as well as through a synthesis of analytical tools (i.e., Non-negative Matrix Factorization, Latent Dirichlet Allocation, and Principal Component Analysis) applied on the findings of our dataset.

Our first step in this exploration (i.e., the descriptive statistics comprehensive mapping) allowed us to discern overarching patterns and trends within the dataset. Notably, a significant majority of the studies originated from the United States. This geographic concentration necessitates a critical evaluation of potential geographical bias as this American-centric focus of the existing research may influence the applicability and generalizability of these findings on a global scale, potentially skewing the perception of music’s effectiveness and its adaptability in different cultural contexts. Recognizing and addressing this limitation is crucial, as the efficacy and implementation of music in pediatric care might exhibit considerable variations across diverse cultural and geographic settings. By the same token, the deployment of quantitative methods in 82% of the reviewed studies marks a significant trend towards prioritizing empirical data. While this highlights the field’s dedication to empirical rigor, it also reveals a notable gap in qualitative research. The scarcity of qualitative studies is a concern as these methodologies can provide valuable context and insights into personal experiences and outcomes, enriching our understanding beyond what can be quantified, as it is also emphasized in the wider medical literature [33]. The underrepresentation of qualitative approaches may lead to an incomplete portrayal of the nuanced impacts of music therapy and music medicine, aspects that quantitative data alone cannot fully capture. This imbalance underscores the need for an integrative research approach that combines both quantitative and qualitative methods, ensuring a more holistic understanding of the role of music in pediatric care.

Expanding upon this analysis, we most importantly observed a pronounced focus within the research on the application of music in medical treatment and therapy, overshadowing its role in development and prevention. This pattern strongly implies that the immediate concerns of treatment and medical therapy are receiving disproportionate attention within the specific research domain. Conversely, areas related to developmental milestones or procedures as well as preventative strategies appear to be less rigorously explored. This discrepancy may signal a prevailing reactive approach in the field, as opposed to a more balanced, proactive strategy that equally values prevention and long-term developmental outcomes. The importance of this finding is also underscored by the current shift in professional medical and clinical bodies advocating for more prevention research and developmental data. For instance, the article by Sabayan et al., [46], titled “Preventive neurology: an emerging field toward brain health” in Neurology, reflects this evolving perspective. Additionally, this observation aligns with the findings of Bielenik et al.’s meta-analysis and review [4], which highlighted a gap in research on long-term outcomes for music therapy with preterm infants.

In conjunction with these observations, our methodological review revealed a striking diversity in data collection methods, ranging from physiological measures to advanced neuroimaging. This diversity exemplifies the field’s openness to various investigative approaches, reflecting a willingness to embrace a range of scientific techniques. However, the broad spectrum of methodologies employed across studies presents a double-edged sword. While this methodological plurality enriches the field, it simultaneously introduces challenges in standardization and comparability of results. The presence of such methodological heterogeneity, as seen in our dataset, underscores the need for consistency in research practices. This finding has been also stressed significantly elsewhere in the same field [28], suggesting that methodological alignment is pivotal for the reliability and replicability of future research, particularly in a field as dynamic and interdisciplinary as music in pediatric care.

Another salient observation from our comprehensive mapping was a strong clinical setting bias, with a significant majority of studies conducted in hospitals. This skew may inadvertently neglect the potential effects of passive music listening in less controlled, everyday environments. Such environments are crucial for a comprehensive understanding of music’s impact on infant development and treatment, especially considering the daily contexts in which children engage with music. Moreover, the representation of professionals within these studies indicated possible regional preferences for certain disciplines, suggesting a need for broader interdisciplinary as well as geographical parallel engagement as stated above. This aspect is critical to avoid academic insularity and to encourage diverse perspectives, enriching in this way the scope and applicability of research findings.

Finally, a notable gap we found in the literature is the lack of detailed reporting on infants’ pathologies and intervention lengths. This omission hampers the ability to synthesize evidence and draw valid conclusions across studies. Therefore, we believe that addressing this gap is essential for enhancing the comparability and robustness of future systematic reviews and for formulating comprehensive, evidence-based recommendations in the field of pediatric care.

In shifting our focus from descriptive statistics to the analytical outcomes based on the findings of these 56 studies, our investigation transitioned from a broad overview to a more nuanced analysis, revealing pivotal trends and insights into the landscape of the passive music listening within pediatric contexts. For example, the dominant exploration of physiological impacts in these studies suggests a strong medical orientation in the field. This focus not only aligns with evidence-based practices crucial for safe and effective pediatric care but also fosters interdisciplinary collaboration. Moreover, it aids in refining pediatric care protocols, attracting necessary funding, and providing quantifiable outcomes for policy-making and educational initiatives in healthcare. However, this focus may also overshadow the potential contributions of interdisciplinary approaches [7], encompassing psychology, neuroscience, and the arts, which could offer a more comprehensive understanding of infant well-being and development.

Moreover, the overwhelming emphasis on pain management in this context raises ethical considerations regarding the suitability of interventions for this vulnerable group. These considerations include ensuring informed consent and child assent, conducting rigorous risk-benefit analyses, preventing potential exploitation of vulnerable participants, monitoring long-term effects, and respecting cultural sensitivities - elements we have already seen missing in the studies of this literature review. Such considerations are crucial to safeguard the welfare and rights of child participants in these studies [2], while they also underscore the necessity for heightened ethical discourse and the establishment of more robust guidelines.

Methodologically, the 56 studies exhibit a strong emphasis on rigorous methodology, reflecting the field’s recognition of its importance. However, this focus might also result in a compartmentalization of approaches, possibly at the expense of more innovative and exploratory research as well as specialization of practice. This is not only evident by the fact that most of the 56 studies focus primarily on immediate physiological impacts - revealing therefore a gap in understanding the long-term implications of these interventions as also mentioned before - but is also indicated by the identification of a monotypic cluster in the Principal Component Analysis (PCA), suggesting that in all of this relevant literature there is only one group of studies heavily reliant on each other, contrary to the expectation of a more diverse range. While this demonstrates a solid line of inquiry for this particular cluster of studies, it could also imply a lack of innovation in both research and practice approaches or subjects, an element that, if evident, has been seen to hinder professional development and individual clinical expertise resulting in turn a suboptimal practice of evidence-based medicine and therapy protocols [34].

On the other hand, this weak but existent centralized approach taken, ranging mostly from pain management to feeding habits, underscores an emerging potential value of narrative synthesis for clinicians. Yet, this method appears underutilized according to our analysis. This is evident in the fragmented nature of reporting across studies, where findings seem to be presented in isolation, focusing narrowly on specific aspects of infant care such as physiological responses, without considering the broader context or interconnections existing between these or other important areas. For instance, research on the impact of music on pain management in infants is reported separately from studies investigating its effects on feeding behaviors (i.e., no actual links were evident throughout our NMF and LDA analysis), missing therefore, in our view, opportunities to explore potential synergies or holistic impacts of music therapy and music medicine in the wider clinical and domestic context. This lack of integrated perspectives in research is further reflected in the disconnect between research findings and their implementation in public policy. Despite overwhelming support for certain interventions, a discernible gap remains between scientific evidence and policy advocacy.

This discrepancy can be traced back to several nuanced elements within our systematic review. For example, we have seen that the studies predominantly rely on quantitative methods, which, while rigorous, may not adequately capture the complexities and contextual variations essential for shaping effective public policies, as evidenced by the limitations highlighted in quantitative health impact assessments [27]. This methodological inclination towards quantitative data often leads to a gap between what is scientifically validated under controlled conditions and what is practically feasible or relevant in the varied scenarios of public health, thereby hindering the transition of these findings into actionable policies. Compounding this issue is the fact that most of the research is conducted in clinical settings, focusing more narrowly on clinical efficacy. This clinical orientation, however, may not fully address the broader considerations necessary for policy development, such as accessibility, cost-effectiveness, and integration into existing health systems, a concern that is also reflected in other conjoining domains in the same context [9]. Policies require a holistic view that encompasses these wider public health perspectives, which seems to be underrepresented in the current research oeuvre. Additionally, the results indicate a lack of emphasis on translational research - studies that bridge the gap between clinical findings and practical application in real-world settings. This absence is significant, as translational research is crucial for understanding how to implement research findings effectively and efficiently in diverse healthcare environments. This absence is significant, as translational research is crucial for understanding how to implement research findings effectively and efficiently in diverse healthcare environments; although admittedly its translation into clinical practice is globally slow [1]. Without this bridge, there remains a chasm between scientific knowledge and its application in public health policy. Lastly, economic evaluations of these interventions are completely lacking, especially concerning the costs and benefits of large-scale implementation. This gap highlights the need for more comprehensive research that encompasses economic aspects, long-term effects, caregiver roles, and policy implications to fully understand and leverage the benefits of music therapy and music medicine in pediatric care, as economic evaluations often remain inaccessible to policymakers [23].

## Conclusion

This systematic review has illuminated the multifaceted role of passive music listening in pediatric care, uncovering both its potential benefits and the gaps in current research. While recognizing the efficacy of music as a therapeutic and developmental tool for infants, there is a compelling need for research that is broader and more culturally diverse, incorporating both qualitative and quantitative methodologies. The future of this field lies in integrating insights from neuroscience with developmental studies, deepening our understanding of how music interacts with the developing brain. Such interdisciplinary research will not only enhance our theoretical knowledge but also improve practical applications in pediatric care worldwide. As we move forward, the challenge is to bridge the gap between scientific understanding and practical application, exploring music’s role beyond clinical settings and into everyday pediatric care. Addressing the ethical dimensions of using music with vulnerable infant populations is also paramount. The aim is to harness the full potential of music therapy and music medicine in a global, culturally nuanced context. By doing so, we can contribute to the holistic well-being and developmental outcomes for children across the globe, ensuring that music becomes a universally accessible and effective tool in pediatric care.

## Data Availability

All data produced in the present study are available upon reasonable request to the authors

## Declarations

### Ethics approval and consent to participate

Not applicable

### Consent for publication

Not applicable

### Availability of data and materials

The datasets used and/or analysed during the current study are available from the corresponding author on reasonable request.

### Authors’ contributions

- Conception: E.P.
- Design of the work: E.P.
- Acquisition, analysis of data: E.P., M.A., R.N.S., E.B., V.P.
- Interpretation of data: E.P., S.E., U.W.V.L, D.H.-A., C.G.
- Have drafted the work or substantively revised it: E.P., M.A., R.N.S.

## Aknowlegments

Not applicable

